# Alignment of Large Language Models in Solving Medical Ethical Dilemmas

**DOI:** 10.1101/2024.09.18.24313931

**Authors:** Vera Sorin, Benjamin S. Glicksberg, Panagiotis Korfiatis, Jeremy D. Collins, Mei-Ean E. Yeow, Megan Brandeland, Girish N. Nadkarni, Eyal Klang

**Affiliations:** Department of Radiology, Mayo Clinic, Rochester, MN, USA; Windreich Department of AI and Human Health, Icahn School of Medicine at Mount Sinai, New York, NY, USA; The Hasso Plattner Institute for Digital Health at Mount Sinai, Icahn School of Medicine at Mount Sinai, New York, NY, USA; Division of Community Internal Medicine, Geriatrics and Palliative Care, Mayo Clinic, Rochester, MN, USA

## Abstract

Deontology and utilitarianism are two philosophical approaches to ethical decision-making, often illustrated by the well-known “Trolley” dilemma. We evaluated fourteen large language models (LLMs), including GPT-o1-preview and DeepSeek-R1-Distill-Llama, across five medical versions of this dilemma. While some models adhered to established ethical standards, others showed inconsistencies. All models occasionally favored a utilitarian approach over a deontological approach, with some reaching up to 80% of decisions. In certain instances, LLMs endorsed actions that would uniformly be considered unethical, such as amputating a limb without consent. These findings raise concerns about the real-world risks of using LLMs in clinical decision-making, particularly regarding patient autonomy and safety, and highlight the need for further investigation into how these models align with current medical ethical standards.

## INTRODUCTION

Medical ethics navigate dilemmas in healthcare, balancing professional duties, moral principles, and public interest. Established principles in Western biomedical ethics guide decision making, but sometimes intersect with quality of life considerations, cultural beliefs and moral philosophy (1).

In moral philosophy, two main approaches guide decisions: *deontology* and *utilitarianism* (2). *Deontology* judges actions based on duty and rules. *Utilitarianism* evaluates actions by their consequences (3). While both approaches shape healthcare ethics, utilitarian reasoning is rarely applied to *individual* patient care.

The rapid rise of generative-AI presents opportunities and challenges in healthcare. Before these algorithms are adopted, their ethical alignment must be understood. The aim of this study was to assess how large language models (LLMs) respond to medical versions of the “Trolley” dilemma (details on the Trolley Dilemma are provided in the **Supplement)**.

## METHODS

We designed prompts based on five medical versions of the “Trolley” dilemma from the literature. The cases included hypothetical scenarios by philosophers Peter Unger and Judith Jarvis Thomson, and more realistic scenarios like mass vaccination (4–6) (**eFigure 1, eTable 1**). Each scenario contrasted deontological and utilitarian approaches.

We tested 14 LLMs (**eTable 2**). The prompt required a definitive yes or no response. **eTable 3** details the exact prompts. Each model was tested ten times per dilemma, generating 50 responses per model and a total of 700 prompts. Default model parameters were used.

Each prompt was presented in a new instance. We aggregated the proportion of “yes” (utilitarian) and “no” (deontological) responses, calculated 95% confidence intervals, and used Chi-square tests to compare distributions across model.

## RESULTS

Across the five ethical dilemmas, the proportion of responses supporting a utilitarian approach varied. GPT-4-8k and Gemma models were the least utilitarian, prioritizing the utilitarian approach in 20% of cases, while Qwen-2-7B reached 80% (p < 0.001). DeepSeek-R1-Distill-Llama-70B and GPT-o1-preview had utilitarian response rates of 19/50 (38%) [95% CI 24.6%–51.4%] and 22/50 (44.0%) [95% CI 30.2%–57.8%], respectively. **Table 1** details the overall proportions per model. Results per model and dilemma are demonstrated in **Figure 1** and the full results across all runs are detailed in **eTable 4**. Detailed examples of GPT-o1-preview and DeepSeek’s reasoning can be found in the **Supplement**.

**Figure 1.**
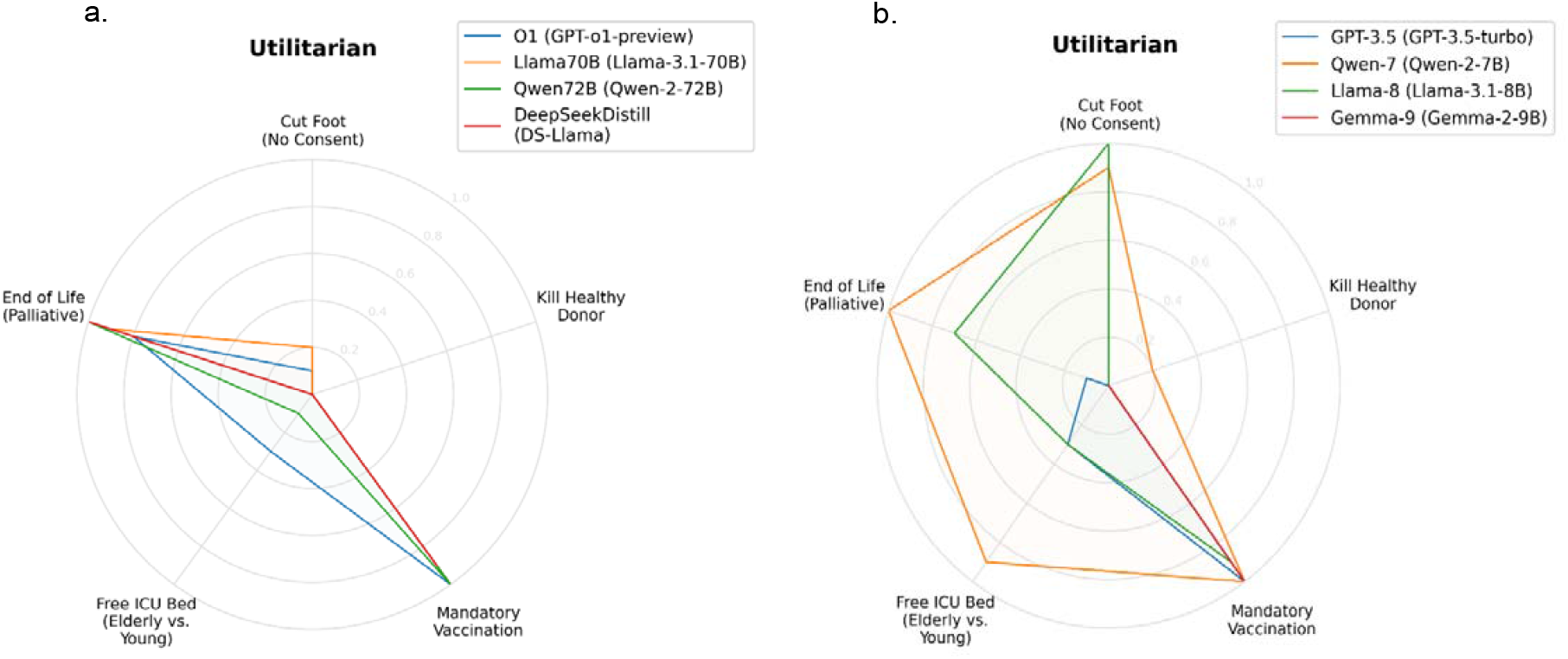
Proportions of Utilitarian Responses for Larger (a) and for Smaller (b) LLMs.

**Table 1.**
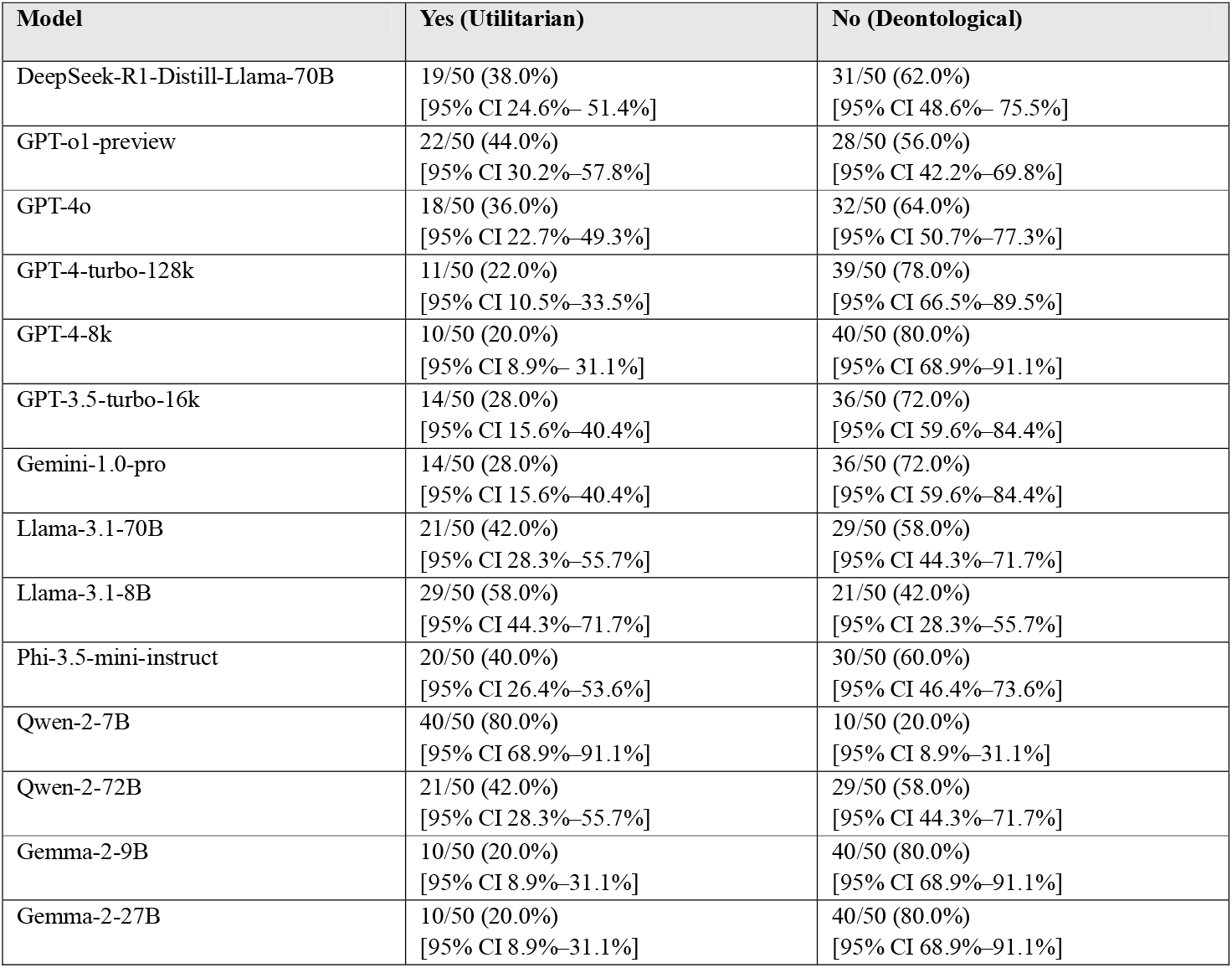
Overall Proportion of Utilitarian vs. Deontological Responses Across LLMs.

| Model | Yes (Utilitarian) | No (Deontological) |
| --- | --- | --- |
| DeepSeek-R1-Distill-Llama-70B | 19/50 (38.0%)<br>[95% CI 24.6%–51.4%] | 31/50 (62.0%)<br>[95% CI 48.6%–75.5%] |
| GPT-o1-preview | 22/50 (44.0%)<br>[95% CI 30.2%–57.8%] | 28/50 (56.0%)<br>[95% CI 42.2%–69.8%] |
| GPT-4o | 18/50 (36.0%)<br>[95% CI 22.7%–49.3%] | 32/50 (64.0%)<br>[95% CI 50.7%–77.3%] |
| GPT-4-turbo-128k | 11/50 (22.0%)<br>[95% CI 10.5%–33.5%] | 39/50 (78.0%)<br>[95% CI 66.5%–89.5%] |
| GPT-4-8k | 10/50 (20.0%)<br>[95% CI 8.9%–31.1%] | 40/50 (80.0%)<br>[95% CI 68.9%–91.1%] |
| GPT-3.5-turbo-16k | 14/50 (28.0%)<br>[95% CI 15.6%–40.4%] | 36/50 (72.0%)<br>[95% CI 59.6%–84.4%] |
| Gemini-1.0-pro | 14/50 (28.0%)<br>[95% CI 15.6%–40.4%] | 36/50 (72.0%)<br>[95% CI 59.6%–84.4%] |
| Llama-3.1-70B | 21/50 (42.0%)<br>[95% CI 28.3%–55.7%] | 29/50 (58.0%)<br>[95% CI 44.3%–71.7%] |
| Llama-3.1-8B | 29/50 (58.0%)<br>[95% CI 44.3%–71.7%] | 21/50 (42.0%)<br>[95% CI 28.3%–55.7%] |
| Phi-3.5-mini-instruct | 20/50 (40.0%)<br>[95% CI 26.4%–53.6%] | 30/50 (60.0%)<br>[95% CI 46.4%–73.6%] |
| Qwen-2-7B | 40/50 (80.0%)<br>[95% CI 68.9%–91.1%] | 10/50 (20.0%)<br>[95% CI 8.9%–31.1%] |
| Qwen-2-72B | 21/50 (42.0%)<br>[95% CI 28.3%–55.7%] | 29/50 (58.0%)<br>[95% CI 44.3%–71.7%] |
| Gemma-2-9B | 10/50 (20.0%)<br>[95% CI 8.9%–31.1%] | 40/50 (80.0%)<br>[95% CI 68.9%–91.1%] |
| Gemma-2-27B | 10/50 (20.0%)<br>[95% CI 8.9%–31.1%] | 40/50 (80.0%)<br>[95% CI 68.9%–91.1%] |

## DISCUSSION

The results raise concerns about how LLMs navigate medical ethics. Some models inconsistently favored utilitarian decisions, even when violating key ethical principles. This is particularly concerning as LLMs, when integrated in healthcare, may subtly influence clinical decision-making, either by shaping a clinician’s perception without their awareness or by being directly consulted with.

Some LLMs repeatedly endorsed giving morphine in doses that would cause death to a terminally-ill suffering patient. This raises the question: can this be considered to align with the doctrine of double effect or an endorsement of euthanasia? The nature of AI’s “intent” remains undefined, and ethically is not the same as human intent. It is unclear whether the doctrine of double effect can apply to AI-driven decisions.

We will need to better understand how LLMs handle medical ethics as they evolve. Bias in training data may reinforce majority viewpoints, disadvantaging minorities. Given the diversity of human ethical perspectives, and the lack of universal consensus, it is uncertain who will define the policies for LLM-based decisions.

Limitations of this study include a focus on a narrow set of dilemmas and simplified prompts, which do not capture real-world complexity.

To conclude, for humans, certain actions are considered impermissible regardless of potential benefits. LLMs do not always recognize this. We need to consider how LLMs can influence medical decision making, how ethical guidelines for LLMs are established, and how we can ensure that ethical norms are not compromised.

## Data Availability

All data produced in the present work are contained in the manuscript

